# Differentials and predictors of hospitalization among the elderly people in India: Evidence from 75^th^ round of National Sample Survey (2017-18)

**DOI:** 10.1101/2021.08.25.21262606

**Authors:** Saddaf Naaz Akhtar, Nandita Saikia

## Abstract

**Introduction:** There are limited evidences on the determinants of hospitalization and its causes in India. We examined the differential in the hospitalization rates and its socio-economic determinants. We also examined the causes of diseases in hospitalization among the elderly (≥60 years) in India.

**Methods:** We used data from 75^th^ round of the National Sample Survey Organizations (NSSO), collected from July 2017 to June 2018. The elderly samples in this survey are 42759, where 11070 were hospitalized, and 31,689 were not hospitalized in the last year or 365 days. We estimated hospitalization rates and carried out binary logistic regression analysis to examine the associations of hospitalization with the background variables. The cause of diseases in hospitalizations were also calculated.

**Results:** Hospitalization rate was lower among female elderly compared to male elderly. Elderly who belongs to middle-old aged groups, non-married, North-Eastern region, Southern region, general caste, health insurance, partially & fully economically dependent elderly have a higher chance of being hospitalized. About 38% elderly were hospitalized due to communicable diseases (CDs), 52% due to non-communicable diseases (NCDs) and 10% due to Injuries & others. Nearly 40% elderly were hospitalized in public hospitals due to CDs, while 52% were hospitalized in private hospitals due to NCDs and 11% due to Injuries & others.

**Conclusions:** Raising awareness, promoting a healthy lifestyle, and improving the quality of good healthcare provisions at the primary level is necessary. Early screening and early treatment for NCDs are needed, which is non-existent in almost all parts of India.

## Introduction

An increasing number of acute hospitalizations of elderly people is seen in many countries ^1–4^. Elderly people need more frequent hospitalization ^5^ because of cardiovascular, respiratory, urinary system, and musculoskeletal diseases ^2^. According to the United Nations Population Fund (UNFPA), India’s aging population stood at 1.36 billion in 2019 due to decreasing fertility rates and improving life expectancy. With increase in life expectancy, there is more concentration of disability and other non-communicable diseases at the old age. As a result, hospitals have to face with a growing number of elderly patients at high risk of adverse events during hospitalization ^6,7^.

Hospitalization is a stressful event and has a high risk of losing autonomy among older adults ^8^ and the oldest-old ^9,10^. Interestingly, it is also an indicator of the demand for restoring health and an integral part of any health system ^11^. The elderly population is subjected to a poor adaption of the health system due to aging and frailty. Several studies have highlighted that older people who are hospitalized due to severe health events, representing a higher health risk ^6,12–14^. With increase in age, the hospitalization outcome turned poorer ^15^. Earlier studies depicted hospitalization has highly complex need because of the multiple co-morbidities, increased functional dependence, physical limitations, and complicated psychosocial problems among elderly ^7^. The significant determinants of hospitalization among elderly were subjected on specific diseases ^16^. Causes of diseases in hospitalization are essential as a public health concern that emphasized treatment and prevention strategies. Injuries and infections among elderly people were highly connected and significant to hospitalizations ^17^.

There have been a limited number of studies based on gender or socio-economic disparities in hospitalization and causes of diseases in hospitalization among elderly. For instance, studies by ^18,19^ have showed that girls have lower chance of hospitalization than boys, especially when households have poor economic status. Higher income quintiles were associated with a higher chance of hospitalization, and the effect is stronger among the elderly males than females in India ^20^. Another study by ^21^ has showed that there is a significant socioeconomic gradient in the distribution of distressed financing with huge disadvantages for marginalized sections, particularly females, elderly and backward social groups. The elderly has a much higher hospitalization rate and has continuing greater socioeconomic differentials in hospitalization rates in India ^11^. Pro-poor inequity exists in public facilities’ use was higher among the elderly than adults in 2014 ^22^. An emerging pattern of high morbidity has been found in Kerala, which required proper treatment and constant medical attention at primary and community levels, especially among elderly people ^23^. A recent study has observed that the bias exists in hospitalizations financing among the elderly females, even the health insurance failed to support elderly females against this gender bias ^24–26^.

Yet, there are limited evidences on the determinants of hospitalization and its causes in India. Studies on predictors of hospitalization and its causes of diseases may help to prioritize effective interventions to achieve equity in hospitalizations and good health. Therefore, the present study examines the differential in the hospitalization rates and it’s socio-economic determinants. The present study also examines the causes of diseases in hospitalisation by background characteristics among the elderly (≥60 years) people in India.

## Methods

### Study design

The present study has used the data from 25^th^ schedule of the 75^th^ round of the National Sample Survey Organizations (NSSO), collected from July 2017 to June 2018. The NSSO is a public organization since 1950 under the Ministry of Statistics and Programme Implementation (MOSPI) of the Government of India. The 75^th^ round of NSSO data is a nationally and state/Union Territory (UT) representative sample of household, cross-sectional, population-based survey.

### Data collection technique

The NSSO collects data on the various issues such as migration, employment, general consumer expenditures, educational attainment, health, Sanitation & hygiene. The 25^th^ schedule of the 75^th^ round of NSSO, known as “Social consumption in Health Survey,” which has been collected information on demographic and socioeconomic factors of the population surveyed. This has mainly focused on health conditions, healthcare financing, insurance coverages by private and public sectors, access to healthcare services, and hospitalization. The hospitalization includes in-patient treatment in one year preceding to the survey and out-patient treatment during 15 days prior to the survey. Hospitalization survey consists of nature of ailments, duration of treatment (days), type of medical institute (level of care), nature of treatments, received any free medical services, total expenditure (including medical and non-medical expenses like medication, consultation and transportation etc.), sources of financing for healthcare expenses. A multistage stratified sampling technique has been adopted in this survey, where 555352 individuals from 113823 households in India. The detailed information about this NSSO are available elsewhere ^27^.

## Analytical sample

The analytical sample comprises twofold: The total sample of elderly information cases was 42759 (excluding two individuals who were self-identified as transgender, including 21902 elderly males & 20857 elderly females). Out of 42759, 11070 were hospitalized, and 31,689 were not hospitalized samples in the last one year or 365 days prior to the survey of NSSO. Also, there were 9282 samples of elderly hospitalized due to various diseases ailments in the last one year or 365 days prior to the survey of NSSO.

### Outcome variable of hospitalization

Three dependent variables were taken in the present study.

∘ The dependent variable is the dichotomous variable of hospitalization. In the questionnaire, it has been asked to the respondent that whether the individual had been hospitalized or not in the last 365 days. *Not hospitalized (no) is taken as “0” and hospitalized (yes) as “1”*.

### Independent variables

The control variables focus on socio-demographic, economic and elderly’s health status, characteristics. These characteristics include the elderly age groups, place of residence, regions, marital status, education level, wealth quintiles, religion, social-groups (caste groups), household members, and health insurance support. The elderly’s health information or characteristics include economic dependency, owned house, physical mobility, living arrangements, state of current perception about health and perception about change in state of health.

### Analysis of cause of hospitalization

We also analyzed the nature of ailment in the last one year or 365 days prior to the survey of NSSO. For each hospitalization, it has been asked to the respondent about the nature of ailments. This nature of ailments has been divided into 63 response categories. The present study has combined these 63 response categories into three main response groups are: *i) Communicable diseases, ii) non-communicable diseases, and iii) injuries & others*. Since the present study is based on elderly, so we have excluded the cases related to maternity and child births.

The hospitalization & treatment characteristics include the duration of stay in the hospital (in days), total health care expenditure (in INR), major sources of healthcare financing and type of hospitalization (level of care) whether hospitalization was at a public hospital or private.

## Statistical Analysis

The primary analysis used univariate, bivariate cross-tabulations and percentages were calculated by socio-demographic, economic, elderly’s health information characteristics. The present study has evaluated the hospitalization rate by background characteristics. Binary logistic regression analysis has been used to examine the associations of hospitalization among elderly in India. Also, the cause of hospitalizations was calculated by socio-demographic, economic, elderly’s health information, hospitalization & treatment characteristics.

## Results

### Characteristics of analytical sample

Table 1 shows the sample size distribution by socio-demographic and economic background characteristics and health information among the elderly in India from 2017-18. About 51% of elderly males and 49% of elderly females—nearly 65% of elderly who belonged to young-old age group. Around 55% elderly was rural residence. Majority of elderly belonged to the Southern region of India. About 48% elderly have no education, 77% of the sample are of Hindu religion and only 21% have insurance coverage support. About 47% were fully economically dependent, 90% of them have owned house and 87% of them reside with other than spouse. Despite that 90% of elderly have physical mobility.

**Table 1.** Sample size distribution of socio-demographic and economic background characteristics and health information among the elderly in India from the period 2017-18 (n=42,759).

| Background Characteristics | Percentage (%) | Number (n) |
| --- | --- | --- |
| **Sex** |  |  |
| Male | 51.22 | 21,902 |
| Female | 48.78 | 20,857 |
| **Age-groups** |  |  |
| Young-old | 64.94 | 27,768 |
| Middle-old | 26.27 | 11,233 |
| Oldest-old | 8.79 | 3,758 |
| **Place of residence** |  |  |
| Rural | 55.19 | 23,597 |
| Urban | 44.81 | 19,162 |
| **Regions** |  |  |
| Northern | 20.57 | 8,794 |
| Nort-Eastern | 9.33 | 3,989 |
| Central | 14.82 | 6,338 |
| Eastern | 16.34 | 6,986 |
| Western | 14.47 | 6,186 |
| Southern | 24.48 | 10,466 |
| **Marital status** |  |  |
| Never* | 31.42 | 13,437 |
| Married | 68.58 | 29,322 |
| **Education** |  |  |
| No education | 48.81 | 20,869 |
| Primary | 28.96 | 12,383 |
| Secondary | 14.33 | 6,128 |
| Higher | 7.9 | 3,379 |
| **Wealth quintiles** |  |  |
| Poorest | 16.82 | 7,191 |
| Poorer | 16.71 | 7,147 |
| Middle | 19.01 | 8,128 |
| Richer | 22.53 | 9,633 |
| Richest | 24.93 | 10,660 |
| **Religion** |  |  |
| Hindus | 77.74 | 33,240 |
| Muslims | 11.54 | 4,934 |
| Others* | 10.72 | 4,585 |
| **Social-group** |  |  |
| Schedule tribes | 9.15 | 3,913 |
| Schedule caste | 14.34 | 6,131 |
| Other backward caste | 38.63 | 16,519 |
| Others (General) | 37.88 | 16,196 |
| **Household members** |  |  |
| Less than 5 | 49.58 | 21,200 |
| More than 5 | 50.42 | 21,559 |
| **Insurance support** |  |  |
| Uncovered | 79.25 | 33,885 |
| Covered | 20.75 | 8,874 |
| **Economic dependency** |  |  |
| Independent | 29.02 | 12,410 |
| Partial dependnet | 23.69 | 10,129 |
| Fully dependent | 47.29 | 20,220 |
| **Owned House** |  |  |
| No | 9.38 | 4,010 |
| Yes | 90.62 | 38,749 |
| **Physical mobility** |  |  |
| Immobile | 1.97 | 844 |
| Partial-mobile | 7.88 | 3,369 |
| physical-mobility | 90.15 | 38,546 |
| **Living arrangements** |  |  |
| Alone* | 1.97 | 841 |
| Only spouse | 10.64 | 4,549 |
| Other than spouse* | 87.39 | 37,369 |
| **State of current perception about health** |  |  |
| Poor | 22.94 | 9,809 |
| Fairly | 68.81 | 29,421 |
| Excellent | 8.25 | 3,529 |
| **Perception about change in state of health** |  |  |
| Worst | 24.35 | 10,412 |
| Nearly same | 56.93 | 24,343 |
| Better than before | 18.72 | 8,004 |
| **Total** | **100** | **42,759** |

**Source :** Authors' own calculation using 75th round of National Sample Survey data.
**Note: 1** Never\*: includes elderly who were never married or separated or divorced. Others\*: includes elderly who were belong to Christians or Sikhs or Jains or Buddhists or others. Alone\*: As an inmate of old age homes or not as an inmate of old age homes. Other than spouse\*: without spouse but with children or other relations or non-relations.

### Hospitalization rates at the national level

Table 2 shows the hospitalization rates of elderly patients who had been hospitalized in the last 365 days by socio-demographic and economic backgrounds characteristics in India, 2017-18. The hospitalization rate among elderly males (7%, p<0.01) is found to be marginally higher than elderly females (6%, p<0.01). With an increase in the elderly age groups, the hospitalization rate gets increases. Southern region of India constitutes the highest elderly hospitalizations. Elderly with richest wealth quintile have highest rate of hospitalization. Nearly 8% elderly who belong to the general caste have the highest hospitalization rate, whereas schedule tribes have the lowest hospitalization rate. Elderly who are fully economically dependent, immobile, and covered with health insurance support have the highest hospitalization rate.

**Table 2.** Hospitalization rates among elderly patients in India by socio-demographic and economic backgrounds characteristics from the period 2017-18 (n=42,759).

| Background Characteristics | Hospitalization rates |
| --- | --- |
| **Sex** |  |
| Male | 7.18 |
| Female | 6.15 |
| **Age-groups** |  |
| Young-old | 5.84 |
| Middle-old | 7.77 |
| Oldest-old | 9.8 |
| **Place of residence** |  |
| Rural | 6.06 |
| Urban | 7.88 |
| **Regions** |  |
| Northern | 6.23 |
| Nort-Eastern | 4.07 |
| Central | 5.15 |
| Eastern | 5.8 |
| Western | 6.37 |
| Southern | 9.11 |
| **Marital status** |  |
| Never* | 6.72 |
| Married | 6.62 |
| **Education** |  |
| No education | 5.44 |
| Primary | 8.43 |
| Secondary | 8.35 |
| Higher | 6.83 |
| **Wealth quintiles** |  |
| Poorest | 4.17 |
| Poorer | 5.56 |
| Middle | 6.97 |
| Richer | 7.72 |
| Richest | 9.11 |
| **Religion** |  |
| Hindus | 6.37 |
| Muslims | 7.39 |
| Others* | 9.37 |
| **Social-group** |  |
| Schedule tribes | 4.61 |
| Schedule caste | 5.61 |
| Other backward caste | 6.47 |
| Others (General) | 7.8 |
| **Household members** |  |
| Less than 5 | 7.31 |
| More than 5 | 5.74 |
| **Insurance support** |  |
| Uncovered | 6.02 |
| Covered | 9.4 |
| **Economic dependency** |  |
| Independent | 5.48 |
| Partial dependnet | 6.82 |
| Fully dependent | 7.33 |
| **Owned House** |  |
| No | 7.34 |
| Yes | 6.59 |
| **Physical mobility** |  |
| Inmobile | 24.71 |
| Partial-mobile | 13.54 |
| physical-mobility | 5.92 |
| **Living arrangements** |  |
| Alone* | 6.47 |
| Only spouse | 8.12 |
| Other than spouse* | 6.41 |
| **State of current perception about health** |  |
| Poor | 12.54 |
| Fairly | 5.52 |
| Excellent | 2.74 |
| **Perception about change in state of health** |  |
| Worst | 12.25 |
| Nearly same | 4.66 |
| Better than before | 6.83 |
| **Total** | **6.66** |

**Source:** Authors' own calculation using 75th round of National Sample Survey data.
**Note: 2** Never\*: includes elderly who were never married or separated or divorced. Others\*: includes elderly who were belong to Christians or Sikhs or Jains or Buddhists or others. Alone\*: As an inmate of old age homes or not as an inmate of old age homes. Other than spouse\*: without spouse but with children or other relations or non-relations. Chi-square test performance shows that there exists statistical significance in the hospitalization of elderly patients at 1 percent and 5 percent level of significance.

### Logistic regression results

Table 3 shows the odds ratios (ORs) results of hospitalization by socio-demographic and economic characteristics of India’s elderly patients from 2017-18. The results indicate that hospitalizations were significantly lower among elderly female than elderly males (OR:0.66, p<0.01). Elderly who belongs to middle-old age groups (OR: 1.08, p<0.01) have higher odds of hospitalization than the young-old age group. Elderly who belongs to North-Eastern region (OR: 1.15, p<0.01) and Southern region (OR: 1.09, p<0.05) are more likely to hospitalized than Northern region.

**Table 3.** Odds ratios (ORs) results of hospitalization, by socio-demographic and economic characteristics of the elderly patients in India from the period 2017-18 (n=42,759).

| **Background Characteristics** | **Odds Ratio [OR]** | **Lower Confidence Intervals [LCI]** | **Upper Confidence Intervals [UCI]** |
| --- | --- | --- | --- |
| **Sex** |  |  |  |
| Male |  |  |  |
| Female | 0.66††† | 0.62 | 0.70 |
| **Age-groups** |  |  |  |
| Young-old |  |  |  |
| Middle-old | 1.08††† | 1.02 | 1.14 |
| Oldest-old | 0.82††† | 0.75 | 0.89 |
| **Place of residence** |  |  |  |
| Rural |  |  |  |
| Urban | 1.04 | 0.98 | 1.10 |
| **Regions** |  |  |  |
| Northern |  |  |  |
| North-Eastern | 1.15††† | 1.04 | 1.27 |
| Central | 1.04 | 0.96 | 1.14 |
| Eastern | 0.88††† | 0.81 | 0.95 |
| Western | 1.04 | 0.96 | 1.13 |
| Southern | 1.09†† | 1.02 | 1.18 |
| **Marital status** |  |  |  |
| Never* |  |  |  |
| Married | 0.83††† | 0.78 | 0.88 |
| **Education** |  |  |  |
| No education |  |  |  |
| Primary | 1.11††† | 1.05 | 1.17 |
| Secondary | 1.12††† | 1.04 | 1.21 |
| Higher | 1.08 | 0.98 | 1.20 |
| **Wealth quintiles** |  |  |  |
| Poorest |  |  |  |
| Poorer | 1.19††† | 1.09 | 1.29 |
| Middle | 1.25††† | 1.15 | 1.36 |
| Richer | 1.34††† | 1.23 | 1.46 |
| Richest | 1.53††† | 1.39 | 1.68 |
| **Religion** |  |  |  |
| Hindus |  |  |  |
| Muslims | 1.05 | 0.98 | 1.14 |
| Others* | 1.00 | 0.92 | 1.08 |
| **Social-group** |  |  |  |
| Schedule tribes |  |  |  |
| Schedule caste | 1.01 | 0.91 | 1.13 |
| Other backward caste | 1.11 | 1.01 | 1.22 |
| Others (General) | 1.15†† | 1.04 | 1.27 |
| **Household members** |  |  |  |
| Less than 5 |  |  |  |
| More than 5 | 0.62††† | 0.59 | 0.65 |
| **Insurance support** |  |  |  |
| Uncovered |  |  |  |
| Covered | 1.23††† | 1.16 | 1.30 |
| **Economic dependency** |  |  |  |
| Independent |  |  |  |
| Partial dependnet | 1.20††† | 1.12 | 1.28 |
| Fully dependent | 1.19††† | 1.11 | 1.27 |
| **Owned House** |  |  |  |
| No |  |  |  |
| Yes | 1.30††† | 1.20 | 1.41 |
| **Physical mobility** |  |  |  |
| Inmobile |  |  |  |
| Partial-mobile | 0.52††† | 0.44 | 0.62 |
| physical-mobility | 0.30††† | 0.26 | 0.35 |
| **Living arrangements** |  |  |  |
| Alone* |  |  |  |
| Only spouse | 0.66††† | 0.56 | 0.77 |
| Other than spouse* | 0.42††† | 0.36 | 0.49 |
| **State of current perception about health** |  |  |  |
| Poor® |  |  |  |
| Fairly | 0.55††† | 0.52 | 0.58 |
| Excellent | 0.23††† | 0.21 | 0.26 |
| **Perception about change in state of health** |  |  |  |
| Worst® |  |  |  |
| Nearly same | 0.52††† | 0.49 | 0.55 |
| Better than before | 0.91††† | 0.84 | 0.98 |

**Source:** Authors' own calculation using 75th round of National Sample Survey data.
**Note: 3** Never\*: includes elderly who were never married or separated or divorced. Others\*: includes elderly who were belong to Christians or Sikhs or Jains or Buddhists or others. Alone\*: As an inmate of old age homes or not as an inmate of old age homes. Other than spouse\*: without spouse but with children or other relations or non-relations. Non-hospitalization is the reference category of the dependent variable; 95% confidence interval in the parentheses; Significant level at: ††† significant at 1 percent and †† significant at 5 percent; ® is the reference category of the independent variables.

Married elderly are significantly less likely to hospitalized compared to never married/divorced/separated (OR: 0.83, p<0.01). With the increase in the educational level and wealth status of elderly the chance of hospitalization increases significantly. Elderly who belongs to general caste (OR: 1.15, p<0.05) are more likely to be hospitalized compared to Schedule tribes (ST) caste. Elderly who opted for health insurance coverage is (OR: 1.23, p<0.01) significantly more likely to be hospitalized than those who did not covered with health insurance. Partially & fully economically dependent elderly and immobile elderly are significantly more likely to be hospitalized. Elderly living with their spouse (OR: 0.66, p<0.01) and other than spouse (OR: 0.42, p<0.01) are less likely to be hospitalized than living alone. Those elderly who has reported that the perception of their current health status is good (OR: 0.55, p<0.01) & excellent (OR: 0.23, p<0.01) and those who reported that no change (OR: 0.52, p<0.01) is seen in their health status are significantly less likely to be hospitalized.

### Causes of diseases in hospitalization among the elderly patents in India

Table 4 shows the causes of diseases in hospitalization by socio-demographic, economic, hospitalization & treatment characteristics of the elderly patients in India from 2017-18. More than 38% of elderly males were hospitalized due to communicable diseases (CDs) and found to be higher than the elderly females. The oldest-old age groups have more than 40% hospitalization due to CDs. Around 10% greater CDs-related hospitalization in rural areas than urban areas. The elderly in India’s Central region have the highest CDs-related hospitalization, whereas the Northern part has the lowest. The elderly with no education has greater hospitalization due to CDs. The poorer elderly has higher hospitalization due to CDs than the rich. The elderly belonging to STs/SCs social group have higher hospitalizations due to CDs than the General caste group. Insured elderly have lower hospitalization due to CDs than uninsured. CDs-related hospitalization is higher among physically mobile than immobile. Elderly who reported excellent about their current health status have higher hospitalization due to CDs compared to the poor perception. Elderly who stayed less than five days & 6-14 days have higher hospitalization due to CDs. CDs-related hospitalization is more when health expenditure is low. Those elderly whose primary source of healthcare financing is from household-income have higher hospitalizations due to CDs. More than 40% of the elderly have preferred public hospitals compared to private hospitals for CDs.

**Table 4.**
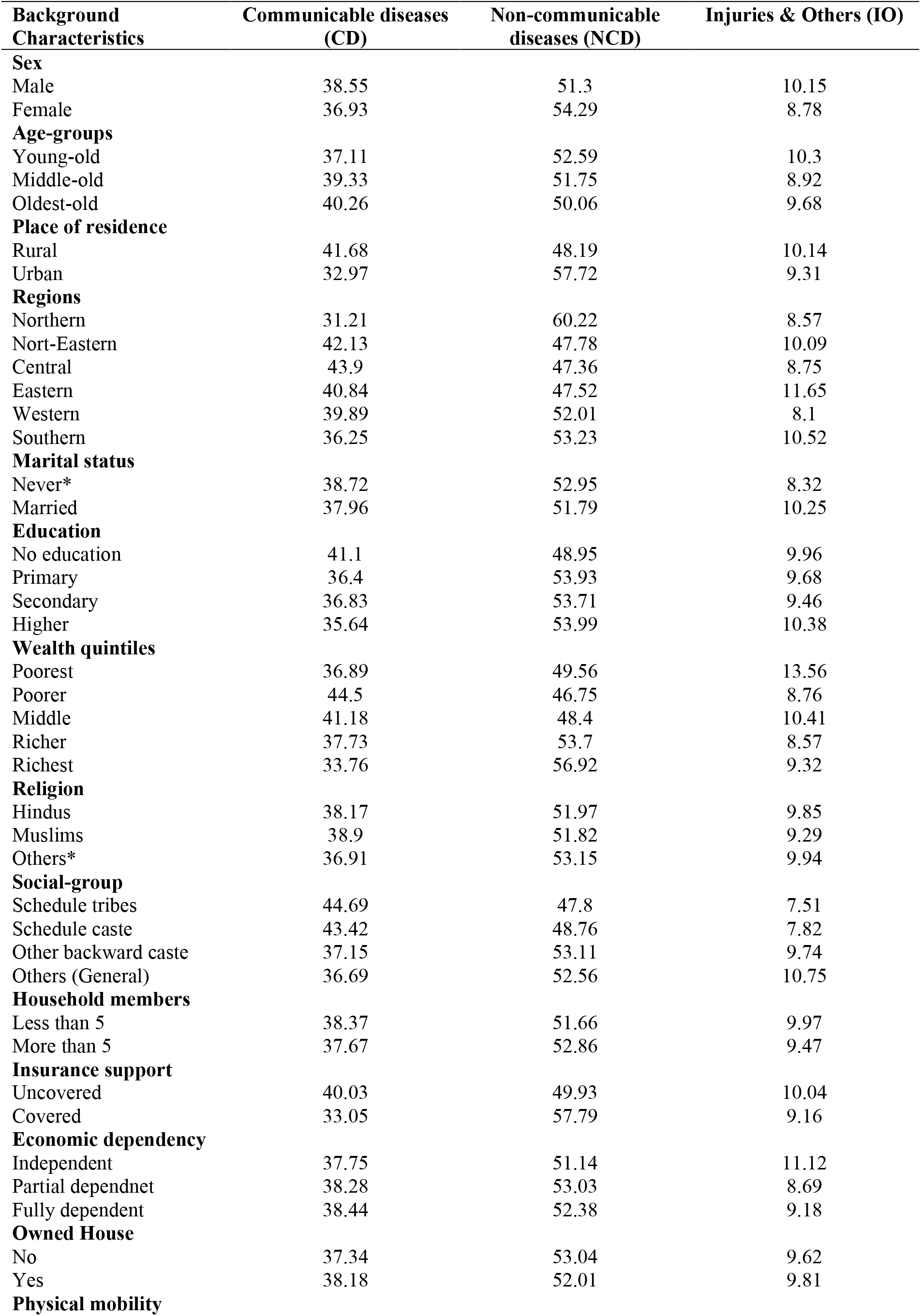
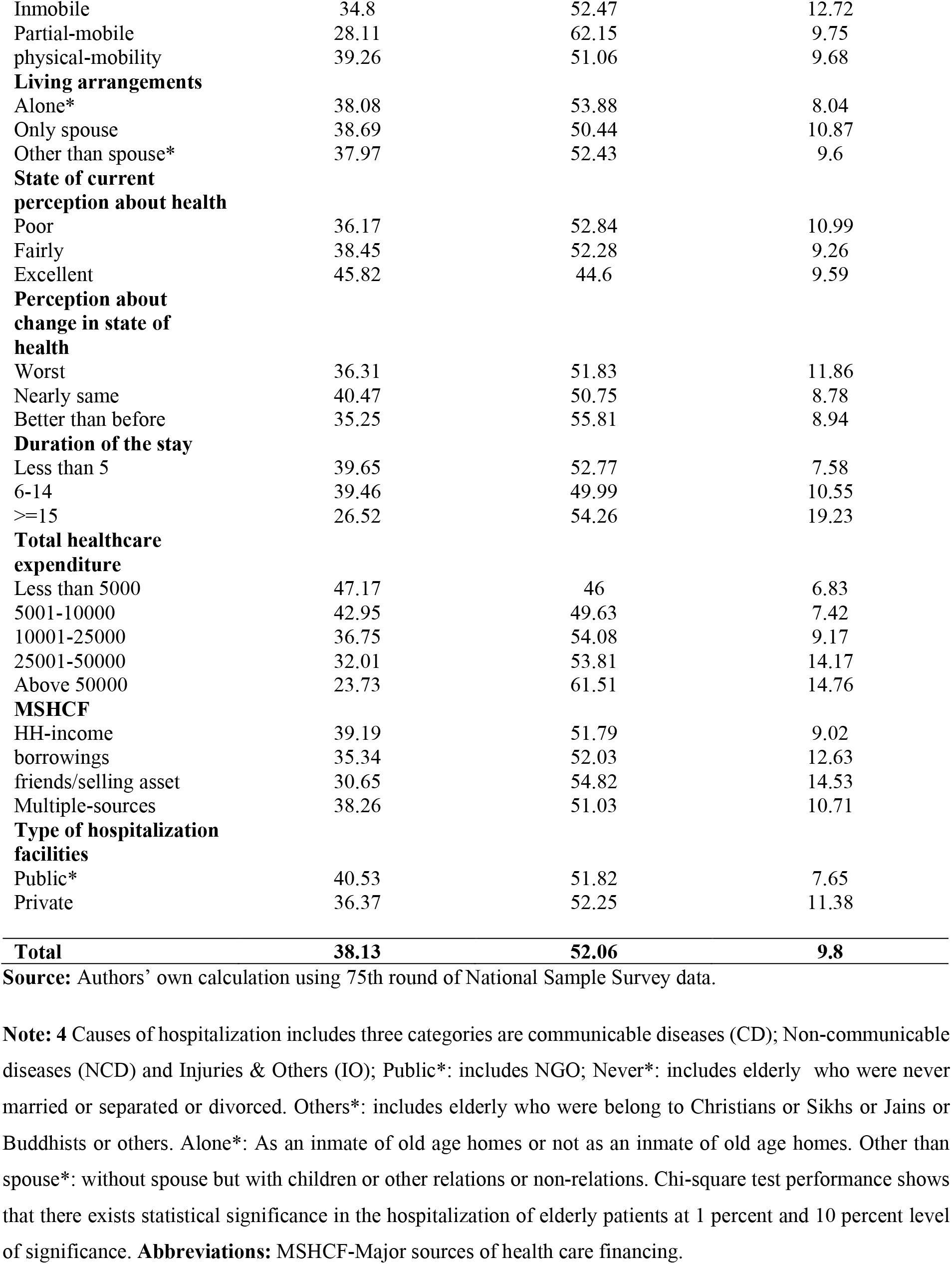
Cause of hospitalization rates among elderly patients in India by socio-demographic and economic backgrounds characteristics from the period 2017-18 (n=9282).

| **Background Characteristics** | **Communicable diseases (CD)** | **Non-communicable diseases (NCD)** | **Injuries & Others (IO)** |
| --- | --- | --- | --- |
| **Sex** |  |  |  |
| Male | 38.55 | 51.3 | 10.15 |
| Female | 36.93 | 54.29 | 8.78 |
| **Age-groups** |  |  |  |
| Young-old | 37.11 | 52.59 | 10.3 |
| Middle-old | 39.33 | 51.75 | 8.92 |
| Oldest-old | 40.26 | 50.06 | 9.68 |
| **Place of residence** |  |  |  |
| Rural | 41.68 | 48.19 | 10.14 |
| Urban | 32.97 | 57.72 | 9.31 |
| **Regions** |  |  |  |
| Northern | 31.21 | 60.22 | 8.57 |
| Nort-Eastern | 42.13 | 47.78 | 10.09 |
| Central | 43.9 | 47.36 | 8.75 |
| Eastern | 40.84 | 47.52 | 11.65 |
| Western | 39.89 | 52.01 | 8.1 |
| Southern | 36.25 | 53.23 | 10.52 |
| **Marital status** |  |  |  |
| Never* | 38.72 | 52.95 | 8.32 |
| Married | 37.96 | 51.79 | 10.25 |
| **Education** |  |  |  |
| No education | 41.1 | 48.95 | 9.96 |
| Primary | 36.4 | 53.93 | 9.68 |
| Secondary | 36.83 | 53.71 | 9.46 |
| Higher | 35.64 | 53.99 | 10.38 |
| **Wealth quintiles** |  |  |  |
| Poorest | 36.89 | 49.56 | 13.56 |
| Poorer | 44.5 | 46.75 | 8.76 |
| Middle | 41.18 | 48.4 | 10.41 |
| Richer | 37.73 | 53.7 | 8.57 |
| Richest | 33.76 | 56.92 | 9.32 |
| **Religion** |  |  |  |
| Hindus | 38.17 | 51.97 | 9.85 |
| Muslims | 38.9 | 51.82 | 9.29 |
| Others* | 36.91 | 53.15 | 9.94 |
| **Social-group** |  |  |  |
| Schedule tribes | 44.69 | 47.8 | 7.51 |
| Schedule caste | 43.42 | 48.76 | 7.82 |
| Other backward caste | 37.15 | 53.11 | 9.74 |
| Others (General) | 36.69 | 52.56 | 10.75 |
| **Household members** |  |  |  |
| Less than 5 | 38.37 | 51.66 | 9.97 |
| More than 5 | 37.67 | 52.86 | 9.47 |
| **Insurance support** |  |  |  |
| Uncovered | 40.03 | 49.93 | 10.04 |
| Covered | 33.05 | 57.79 | 9.16 |
| **Economic dependency** |  |  |  |
| Independent | 37.75 | 51.14 | 11.12 |
| Partial dependnet | 38.28 | 53.03 | 8.69 |
| Fully dependent | 38.44 | 52.38 | 9.18 |
| **Owned House** |  |  |  |
| No | 37.34 | 53.04 | 9.62 |
| Yes | 38.18 | 52.01 | 9.81 |
| **Physical mobility** |  |  |  |
| Inmobile | 34.8 | 52.47 | 12.72 |
| Partial-mobile | 28.11 | 62.15 | 9.75 |
| physical-mobility | 39.26 | 51.06 | 9.68 |
| **Living arrangements** |  |  |  |
| Alone* | 38.08 | 53.88 | 8.04 |
| Only spouse | 38.69 | 50.44 | 10.87 |
| Other than spouse* | 37.97 | 52.43 | 9.6 |
| **State of current perception about health** |  |  |  |
| Poor | 36.17 | 52.84 | 10.99 |
| Fairly | 38.45 | 52.28 | 9.26 |
| Excellent | 45.82 | 44.6 | 9.59 |
| **Perception about change in state of health** |  |  |  |
| Worst | 36.31 | 51.83 | 11.86 |
| Nearly same | 40.47 | 50.75 | 8.78 |
| Better than before | 35.25 | 55.81 | 8.94 |
| **Duration of the stay** |  |  |  |
| Less than 5 | 39.65 | 52.77 | 7.58 |
| 6-14 | 39.46 | 49.99 | 10.55 |
| >=15 | 26.52 | 54.26 | 19.23 |
| **Total healthcare expenditure** |  |  |  |
| Less than 5000 | 47.17 | 46 | 6.83 |
| 5001-10000 | 42.95 | 49.63 | 7.42 |
| 10001-25000 | 36.75 | 54.08 | 9.17 |
| 25001-50000 | 32.01 | 53.81 | 14.17 |
| Above 50000 | 23.73 | 61.51 | 14.76 |
| **MSHCF** |  |  |  |
| HH-income | 39.19 | 51.79 | 9.02 |
| borrowings | 35.34 | 52.03 | 12.63 |
| friends/selling asset | 30.65 | 54.82 | 14.53 |
| Multiple-sources | 38.26 | 51.03 | 10.71 |
| **Type of hospitalization facilities** |  |  |  |
| Public* | 40.53 | 51.82 | 7.65 |
| Private | 36.37 | 52.25 | 11.38 |
| **Total** | **38.13** | **52.06** | **9.8** |

**Source:** Authors' own calculation using 75th round of National Sample Survey data.
**Note: 4** Causes of hospitalization includes three categories are communicable diseases (CD); Non-communicable diseases (NCD) and Injuries & Others (IO); Public\*: includes NGO; Never\*: includes elderly who were never married or separated or divorced. Others\*: includes elderly who were belong to Christians or Sikhs or Jains or Buddhists or others. Alone\*: As an inmate of old age homes or not as an inmate of old age homes. Other than spouse\*: without spouse but with children or other relations or non-relations. Chi-square test performance shows that there exists statistical significance in the hospitalization of elderly patients at 1 percent and 10 percent level of significance. **Abbreviations:** MSHCF-Major sources of health care financing.

Nearly 54% of the elderly females were hospitalized due to non-communicable diseases, significantly higher than elderly males. About 53% of young-old age were hospitalized due to NCDs. The highest hospitalizations due to NCDs were found in the Northern regions (60%) and urban areas. With the increase in educational level, the hospitalization due to NCDs increases. NCDs-related hospitalizations are higher among the richest & richer, which are completely opposite in CDs-related hospitalizations. There has not much difference by religion but by caste groups where OBCs & General caste has higher NCDs-related hospitalizations than STs/SCs. Despite that, NCDs-related hospitalization is high among elderly who were covered with health insurance. Poor perception of current health status and better than before perception about change in health status have higher NCDs-related hospitalization. NCDs-related hospitalization is higher when the duration of stay is high. The elderly who are partially dependent and partial mobile were found to have higher hospitalization due to NCDs. The elderly, whose primary source of healthcare financing is from friends/selling assets, has higher hospitalizations due to NCDs. NCDs-related hospitalization is substantially higher when total healthcare expenditure is high.

However, elderly males have more hospitalization than elderly females due to injuries & others ailments. Around 10% of the elderly in rural areas have higher hospitalization due to Injuries & Others than urban areas. The elderly belonged to the Eastern region, higher education, and poorest wealth quintiles have shown the highest hospitalization due to Injuries & Others. Injuries & Others-related hospitalization were higher among the General caste compared to STs/SCs. The elderly who are economically independent and immobile have significantly greater hospitalization due to Injuries & Others. Elderly living only with a spouse were more hospitalized due to Injuries & Others than living alone. Injuries & Others-related hospitalization was higher among the elderly who have the worst perception of change in health. Injuries & Others-related hospitalization are substantially higher when total healthcare expenditure is high. The elderly, whose primary source of healthcare financing is from friends/selling assets, has higher hospitalizations due to Injuries & Others. The majority of the elderly have preferred private hospitals for hospitalizations due to Injuries & Others compared to public hospitals.

## Discussions

Our study has revealed that the differential exists in the hospitalization and cause of diseases in hospitalization among the elderly people in India. Our findings suggest that the overall hospitalization rates were lesser among the Indian elderly than in other countries like United States ^28^, Brazil ^29,30^, China ^31^. The finding of this study has confirmed from the previous studies; old age people has been found to be higher hospitalization rate ^11,17,32–35^ due to non-communicable, which may be due to the timely policy formulation given the population ageing and drastic shifting of disease from communicable to non-communicable diseases ^36,37^.

Our study confirmed that the elderly living in urban residence and elderly males have shown higher hospitalization rate where these findings were consistent with previous research ^11,17,32– 35,38^. Our results revealed that elderly women have lower odds of hospitalization than the elderly men and similar patterns of hospitalizations were also observed in other countries like-England ^39^, Norway ^17,40^, Italy ^41^, but countries like Denmark ^42^, Finnland ^40^, South Korea ^43^ and Brazil ^44^ have shown higher hospitalization among older women than older men.

Interestingly, our study have showed that married elderly was found to be less likely to be hospitalized than unmarried, these finding are analogous to the other previous studies ^11,35,45– 48^. Our study also confirmed that elderly living alone are more likely to be hospitalized than living with spouse or other than spouse and were also documented in other studies ^35,49–53^.

Further, our study has revealed that the elderly belonging to the North-Eastern region & Southern region of India were more likely to be hospitalized than Northern region. Yet, there has not been any literature which supports the high hospitalization in North-Eastern region. Though, there has been plenty of research which shows higher hospitalization in Southern region ^11,32,33^. It may be due to the larger share of the elderly population were seen in the Southern region in India ^54,55^, and greater treatment-seeking behaviors ^32,56^ because of the availability and accessibility of geriatric healthcare facilities. Our finding has also highlighted that the elderly who were covered with health insurance support were significantly more likely to get hospitalized in India, and were also reflected in other studies ^35,57–59^. Elderly who was fully economical dependent and immobile have highest hospitalization rate which have been also documented in previous studies. Literature suggests that negative self-perceptions of health were associated with greater chance of hospitalization among elderly in Brazil^60,61^ which also supports our finding.

Though ST/SCs are officially designated disadvantaged social groups in India, previous study^22^ also showed that they have higher hospitalizations in 1995 and lower hospitalization in 2014. Our recent finding shows that the General caste social group has a higher chance of hospitalization. Apart from that, our results has found that increase in wealth quintiles and educational attainment increases the chances the hospitalization and vice-versa and can also be seen in earlier studies^11,62–65^.

Notwithstanding, the burden of hospitalization due to NCDs among elderly are showing rising trend globally as well as in India ^11,63,64,66–72^. Our finding showed that young-old and elderly females were more hospitalized due to NCDs, so as in other studies ^11,68,73^; conversely, earlier research has also found that elderly males have higher hospitalizations due to NCDs ^72^. Elderly belong to Northern region have highest hospitalizations due to NCDs which were also seen in past research ^70,73,74^. Higher hospitalization rates due to NCDs among disadvantage social-group exists in Italy ^62^, Sweden ^75,76^, United States ^77^, Australia ^78^, New Zealand ^79^, so it is seen in India where elderly belonging to OBC were prone to hospital admission due to NCDs. Despite that, primary sources of healthcare financing from friends/selling assets have higher hospitalizations due to NCDs, and other studies have shown that sources of hospitalization financing were determined by the household’s decision to seek treatment ^19,24,25,80^. Majority of the elderly has preferred private hospitals for NCDs and Injuries & Others compared to public hospitals and earlier studies have found the similar results ^81^, this may be due to the poor accessibility of health institutions in public hospital ^82^. Still, people prefer to avail health care services from private health centers irrespective of their financial status ^83^.

## Limitations

The present study has limitations: First, the NSSO data is a cross-sectional survey due to which temporal ambiguity averted to draw causal inferences based on our findings. Secondly, other significant factors can also predict hospitalizations and provide insightful results, such as lifestyle factors, behavioral factors, obesity, mental state, and several personal habits like smoking cigarettes, drinking alcohol, consuming tobacco, or other harmful substances. But this information was not available in our study. Lastly, respondents belonging to higher wealth quintiles were highly reported which may be due to their higher level of health consciousness reporting-but this is a risk of under reporting. Even with these limitations, the hospitalization issues among the elderly are beneficial to understand the current circumstances of CDs, NCDs and injury & Other diseases for India and its states to formulate health policy.

## Conclusion

As the elderly population is on the rise in India, there is an increasing trend in hospitalization due to NCDs. Therefore, there is a need to improve the critical healthcare services among the elderly in India. Raising awareness, promotion of healthy life style and improving the quality of good healthcare provisions at primary level is a necessity. At the same time, early screening and early treatment for NCDs are needed, which is non-existent in almost all parts of India. It is essential to necessitate and identify the important factors that best predict hospitalization or re-visit of hospital admission. This can provide a way forward for the health policymakers to potentially alter the future research in order to reduce associated comorbidities, unnecessary hospitalizations and other medical complications. Although, the medical advances in India has made rapid strides in the past few decades, it is burdened none the less since the doctor-patient ratio is very low. It is important to develop preventive measures in order to minimize the accidents and causalities to avoid substantial cost associated with the health care elderly.

## Data Availability

The data analyzed are publicly available.

http://mospi.nic.in/unit-level-data-report-nss-75th-round-july-2017-june-2018-schedule-250social-consumption-health

## Declarations

### Ethics approval and consent to participate

Not applicable.

### Consent for publication

Not applicable.

### Availability of data and materials

The data analyzed are publicly available. http://mospi.nic.in/unit-level-data-report-nss-75th-round-july-2017-june-2018-schedule-250social-consumption-health

## Acknowledgement

None

## References

1. Campbell SE, Seymour DG, Primrose WR, Lynch JE, Dunstan E, Espallargues M, et al. A multi-centre European study of factors affecting the discharge destination of older people admitted to hospital: analysis of in-hospital data from the ACMEplus project. Age and Ageing [Internet]. 2005 Sep 1 [cited 2020 Sep 11];34(5):467–75. Available from: http://academic.oup.com/ageing/article/34/5/467/40349/A-multicentre-European-study-of-factors-affecting

2. Kabaday F, Tokgoz F. Hospitalization and Mortality Rates in Patients with Respiratory Diseases in the Very Elderly Population. J Gerontol Geriatr Res [Internet]. 2016 [cited 2021 Feb 19];05(02). Available from: http://www.omicsgroup.org/journals/hospitalization-and-mortality-rates-in-patients-with-respiratory-diseasesin-the-very-elderly-population-2167-7182-1000293.php?aid=71650

3. Masotti L, Ceccarelli E, Cappelli R, Barabesi L, Guerrini M, Forconi S. Length of hospitalization in elderly patients with community-acquired pneumonia. Aging Clin Exp Res [Internet]. 2000 Feb [cited 2021 Jun 30];12(1):35–41. Available from: http://link.springer.com/10.1007/BF03339826

4. Silva TJA, Jerussalmy CS, Farfel JM, Curiati JAE, Jacob-Filho W. Predictors of in-hospital mortality among older patients. Clinics [Internet]. 2009 [cited 2020 Sep 10];64(7):613–8. Available from: http://www.scielo.br/scielo.php?script=sci_arttext&pid=S1807-59322009000700002&lng=en&nrm=iso&tlng=en

5. Neouze A, Dechartres A, Legrain S, Raynaud-Simon A, Gaubert-Dahan M-L, Bonnet-Zamponi D. Hospitalization of elderly in an Acute-Care Geriatric Department. Gériatrie et Psychologie Neuropsychiatrie du Viellissement [Internet]. 2012 Jun [cited 2021 Jan 20];10(2):143–50. Available from: http://www.john-libbey-eurotext.fr/medline.md?doi=10.1684/pnv.2012.0350

6. Gronewold J, Dahlmann C, Jäger M, Hermann DM. Identification of hospitalized elderly patients at risk for adverse in-hospital outcomes in a university orthopedics and trauma surgery environment. Kou YR, editor. PLoS ONE [Internet]. 2017 Nov 10 [cited 2021 May 1];12(11):e0187801. Available from: https://dx.plos.org/10.1371/journal.pone.0187801

7. Singh I. Assessment and Management of Older People in the General Hospital Setting. In: Zawada ET, editor. Challenges in Elder Care [Internet]. InTech; 2016 [cited 2021 Jun 24]. Available from: http://www.intechopen.com/books/challenges-in-elder-care/assessment-and-management-of-older-people-in-the-general-hospital-setting

8. Rossi AP, Rubele S, Pelizzari L, Fantin F, Morgante S, Marchi O, et al. Hospitalization Effects on Physical Performance and Muscle Strength in Hospitalized Elderly Subjects. J Gerontol Geriatr Res [Internet]. 2017 [cited 2021 Jan 27];06(02). Available from: https://www.omicsgroup.org/journals/hospitalization-effects-on-physical-performance-and-muscle-strength-in-hospitalized-elderly-subjects-2167-7182-1000401.php?aid=86957

9. Creditor MC. Hazards of Hospitalization of the Elderly. Annals of Internal Medicine [Internet]. 1993 [cited 2021 Jan 20];(118):219–23. Available from: http://www.drsharonsee.com/uploads/5/9/6/6/59668435/hazardshospitaliz.pdf

10. Hoenig HM, Rubenstein LZ. Hospital-Associated Deconditioning and Dysfunction. Journal of the American Geriatrics Society [Internet]. 1991 Feb [cited 2021 Jan 20];39(2):220–2. Available from: http://doi.wiley.com/10.1111/j.1532-5415.1991.tb01632.x

11. Pandey A, Ploubidis GB, Clarke L, Dandona L. Hospitalisation trends in India from serial cross-sectional nationwide surveys: 1995 to 2014. BMJ Open [Internet]. 2017 Dec [cited 2021 Feb 22];7(12):e014188. Available from: https://bmjopen.bmj.com/lookup/doi/10.1136/bmjopen-2016-014188

12. Covinsky KE, Palmer RM, Fortinsky RH, Counsell SR, Stewart AL, Kresevic D, et al. Loss of Independence in Activities of Daily Living in Older Adults Hospitalized with Medical Illnesses: Increased Vulnerability with Age. Journal of the American Geriatrics Society [Internet]. 2003 Apr [cited 2021 Jan 20];51(4):451–8. Available from: http://doi.wiley.com/10.1046/j.1532-5415.2003.51152.x

13. Dendukuri N, McCusker J, Belzile E. The Identification of Seniors At Risk Screening Tool: Further Evidence of Concurrent and Predictive Validity: NEW RESULTS ON THE VALIDITY OF THE ISAR SCALE. Journal of the American Geriatrics Society [Internet]. 2004 Feb [cited 2021 May 1];52(2):290–6. Available from: http://doi.wiley.com/10.1111/j.1532-5415.2004.52073.x

14. Prasad S. Does Hospitalization Make Elderly Households Poor? An Examination of the Case of Kerala, India. Social Policy & Admin [Internet]. 2007 Aug [cited 2021 Jan 27];41(4):355–71. Available from: http://doi.wiley.com/10.1111/j.1467-9515.2007.00558.x

15. Bakerjuan D. Hospital Care and Older Adults [Internet]. 2020. Available from: https://www.msdmanuals.com/professional/geriatrics/providing-care-to-older-adults/hospital-care-and-older-adults

16. Liotta G, Gilardi F, Orlando S, Rocco G, Proietti MG, Asta F, et al. Cost of hospital care for the older adults according to their level of frailty. A cohort study in the Lazio region, Italy. Ciccozzi M, editor. PLoS ONE [Internet]. 2019 Jun 11 [cited 2021 Feb 20];14(6):e0217829. Available from: https://dx.plos.org/10.1371/journal.pone.0217829

17. Gjestsen MT, Brønnick K, Testad I. Characteristics and predictors for hospitalizations of home-dwelling older persons receiving community care: a cohort study from Norway. BMC Geriatr [Internet]. 2018 Dec [cited 2021 Jan 20];18(1):203. Available from: https://bmcgeriatr.biomedcentral.com/articles/10.1186/s12877-018-0887-z

18. Asfaw A, Klasen S, Lamanna F. Intra-household Gender Disparities in Childrens Medical Care before Death in India. In: Proceedings of the German Development Economics Conference, Zürich 2008, No 21 [Internet]. Zürich; 2007. (IZA Discussion Paper No. 2586). Available from: https://www.econstor.eu/bitstream/10419/39911/1/AEL_2008_21_klasen.pdf

19. Asfaw A, Lamanna F, Klasen S. Gender gap in parents’ financing strategy for hospitalization of their children: evidence from India. Health Econ [Internet]. 2010 Mar [cited 2021 Feb 22];19(3):265–79. Available from: http://doi.wiley.com/10.1002/hec.1468

20. Roy K, Chaudhuri A. Influence of socioeconomic status, wealth and financial empowerment on gender differences in health and healthcare utilization in later life: evidence from India. Social Science & Medicine [Internet]. 2008 May [cited 2021 May 15];66(9):1951–62. Available from: https://linkinghub.elsevier.com/retrieve/pii/S0277953608000415

21. Joe W. Distressed financing of household out-of-pocket health care payments in India: incidence and correlates. Health Policy and Planning [Internet]. 2015 Jul 1 [cited 2021 Feb 22];30(6):728–41. Available from: https://academic.oup.com/heapol/article-lookup/doi/10.1093/heapol/czu050

22. Pandey A, Ploubidis GB, Clarke L, Dandona L. Horizontal inequity in outpatient care use and untreated morbidity: evidence from nationwide surveys in India between 1995 and 2014. Health Policy and Planning [Internet]. 2017 Sep 1 [cited 2021 Feb 22];32(7):969–79. Available from: https://academic.oup.com/heapol/article/32/7/969/3737837

23. Paul K, Chaudhary M, Chowdhary R, Sengupta R. Changes in levels of morbidity and hospitalisation in Kerala: A district level analysis (1995–2014). Clinical Epidemiology and Global Health [Internet]. 2020 Mar [cited 2021 Jun 30];8(1):21–8. Available from: https://linkinghub.elsevier.com/retrieve/pii/S2213398419300442

24. Kumar K, Singh A, James KS, McDougal L, Raj A. Gender bias in hospitalization financing from borrowings, selling of assets, contribution from relatives or friends in India. Social Science & Medicine [Internet]. 2020 Sep [cited 2021 Feb 22];260:113222. Available from: https://linkinghub.elsevier.com/retrieve/pii/S027795362030441X

25. Moradhvaj, Saikia N. Gender disparities in health care expenditures and financing strategies (HCFS) for inpatient care in India. SSM - Population Health [Internet]. 2019 Dec [cited 2021 Feb 22];9:100372. Available from: https://linkinghub.elsevier.com/retrieve/pii/S2352827318302787

26. Saikia N, Moradhvaj Bora JK. Gender Difference in Health-Care Expenditure: Evidence from India Human Development Survey. Sen U, editor. PLoS ONE [Internet]. 2016 Jul 8 [cited 2021 Aug 19];11(7):e0158332. Available from: https://dx.plos.org/10.1371/journal.pone.0158332

27. Government of India, Ministry of Statistics and Programme Implementation. Unit Level data & Report on NSS 75th Round for Schedule-25.0, July 2017-June 2018, (Social Consumption: Health) [Internet]. 2020 [cited 2021 Jan 20]. Available from: https://mospi.gov.in/web/mospi/download-tables-data/-/reports/view/templateTwo/16202?q=TBDCAT

28. Statista. Percentage of U.S. population with a hospitalization in past year from 2000 to 2018, by age [Internet]. 2021 [cited 2021 Aug 19]. Available from: https://www.statista.com/statistics/184447/us-population-with-a-hospitalization-by-age/

29. Cordeiro P, Martins M. Mortalidade hospitalar em pacientes idosos no Sistema عnico de Saúde, região Sudeste. Rev saúde pública [Internet]. 2018 Jul 20 [cited 2021 Aug 19];52:69. Available from: https://www.revistas.usp.br/rsp/article/view/148323

30. Zhao Q, Coelho MSZS, Li S, Saldiva PHN, Abramson MJ, Huxley RR, et al. Trends in Hospital Admission Rates and Associated Direct Healthcare Costs in Brazil: A Nationwide Retrospective Study between 2000 and 2015. The Innovation [Internet]. 2020 May [cited 2021 Aug 19];1(1):100013. Available from: https://linkinghub.elsevier.com/retrieve/pii/S2666675820300138

31. Ma S, Zhou X, Jiang M, Li Q, Gao C, Cao W, et al. Comparison of access to health services among urban-to-urban and rural-to-urban older migrants, and urban and rural older permanent residents in Zhejiang Province, China: a cross-sectional survey. BMC Geriatr [Internet]. 2018 Dec [cited 2021 Aug 19];18(1):174. Available from: https://bmcgeriatr.biomedcentral.com/articles/10.1186/s12877-018-0866-4

32. Dilip TR. Understanding levels of morbidity and hospitalization in Kerala, India. Bulletin of the World Health Organization : the International Journal of Public Health [Internet]. 2002;80(9):746–51. Available from: https://apps.who.int/iris/handle/10665/71532

33. Moneer Alam, Anup Karan. Elderly Health In India: Dimension, Differentiation and Determination [Internet]. UNFPA; 2011 [cited 2021 Jun 30]. Available from: http://rgdoi.net/10.13140/2.1.4232.5128

34. Patel R, Chauhan S. Gender differential in health care utilisation in India. Clinical Epidemiology and Global Health [Internet]. 2020 Jun [cited 2021 Jul 1];8(2):526–30. Available from: https://linkinghub.elsevier.com/retrieve/pii/S2213398419304154

35. Ranjan A, Muraleedharan VR. Equity and elderly health in India: reflections from 75th round National Sample Survey, 2017–18, amidst the COVID-19 pandemic. Global Health [Internet]. 2020 Dec [cited 2021 Feb 22];16(1):93. Available from: https://globalizationandhealth.biomedcentral.com/articles/10.1186/s12992-020-00619-7

36. Nath A, Ingle G. Geriatric health in India: Concerns and solutions. Indian J Community Med [Internet]. 2008 [cited 2021 Jun 30];33(4):214. Available from: http://www.ijcm.org.in/text.asp?2008/33/4/214/43225

37. Tripathy JP. Geriatric care in India: A long way to go. J Midlife Health. 2014 Oct;5(4):205–6.

38. Olofsson M, Jansson J-H, Boman K. Predictors for hospitalizations in elderly patients with clinical symptoms of heart failure: A 10-year observational primary healthcare study. Journal of Clinical Gerontology and Geriatrics [Internet]. 2016 Jun [cited 2021 Feb 19];7(2):53–9. Available from: https://linkinghub.elsevier.com/retrieve/pii/S2210833516000046

39. Simmonds SJ, Syddall HE, Walsh B, Evandrou M, Dennison EM, Cooper C, et al. Understanding NHS hospital admissions in England: linkage of Hospital Episode Statistics to the Hertfordshire Cohort Study. Age and Ageing [Internet]. 2014 Sep [cited 2021 Jul 26];43(5):653–60. Available from: https://academic.oup.com/ageing/article-lookup/doi/10.1093/ageing/afu020

40. Suominen-Taipale AL, Martelin T, Koskinen S, Holmen J, Johnsen R. Gender differences in health care use among the elderly population in areas of Norway and Finland. A cross-sectional analysis based on the HUNT study and the FINRISK Senior Survey. BMC Health Serv Res [Internet]. 2006 Dec [cited 2021 Jul 26];6(1):110. Available from: https://bmchealthservres.biomedcentral.com/articles/10.1186/1472-6963-6-110

41. Rozzini R, Sleiman I, Maggi S, Noale M, Trabucchi M. Gender Differences and Health Status in Old and Very Old Patients. Journal of the American Medical Directors Association [Internet]. 2009 Oct [cited 2021 Jul 26];10(8):554–8. Available from: https://linkinghub.elsevier.com/retrieve/pii/S1525861009001340

42. Höhn A, Oksuzyan A, Lindahl-Jacobsen R, Christensen K, Seaman R. Gender differences in time to first hospital admission at age 60 in Denmark, 1995–2014. Eur J Ageing [Internet]. 2021 Mar 27 [cited 2021 Jul 26]; Available from: http://link.springer.com/10.1007/s10433-021-00614-w

43. Son Y, Won MH. Gender differences in the impact of health literacy on hospital readmission among older heart failure patients: A prospective cohort study. J Adv Nurs [Internet]. 2020 Jun [cited 2021 Jul 26];76(6):1345–54. Available from: https://onlinelibrary.wiley.com/doi/10.1111/jan.14328

44. Barreto SM, Kalache A, Giatti L. Does health status explain gender dissimilarity in healthcare use among older adults? Cad Saúde Pública [Internet]. 2006 Feb [cited 2021 Jul 26];22(2):347–55. Available from: http://www.scielo.br/scielo.php?script=sci_arttext&pid=S0102-311X2006000200012&lng=en&tlng=en

45. Butler JR, Morgan M. Marital status and hospital use. Journal of Epidemiology & Community Health [Internet]. 1977 Sep 1 [cited 2021 Jul 26];31(3):192–8. Available from: https://jech.bmj.com/lookup/doi/10.1136/jech.31.3.192

46. Gordon HS, Rosenthal GE. Impact of marital status on outcomes in hospitalized patients. Evidence from an academic medical center. Arch Intern Med. 1995 Dec 11;155(22):2465–71.

47. Metersky ML, Fine MJ, Mortensen EM. The Effect of Marital Status on the Presentation and Outcomes of Elderly Male Veterans Hospitalized for Pneumonia. Chest [Internet]. 2012 Oct [cited 2021 Jul 26];142(4):982–7. Available from: https://linkinghub.elsevier.com/retrieve/pii/S0012369212605686

48. Hayes RM, Carter PR, Gollop ND, Reynolds J, Uppal H, Sarma J, et al. 108 The Impact of Marital Status on Mortality and Length of Stay in Patients Admitted with Myocardial Infarction: Abstract 108 Table 1. Heart [Internet]. 2016 Jun [cited 2021 Jul 26];102(Suppl 6):A77.1-A77. Available from: https://heart.bmj.com/lookup/doi/10.1136/heartjnl-2016-309890.108

49. Mahoney JE, Eisner J, Havighurst T, Gray S, Palta M. Problems of older adults living alone after hospitalization. J Gen Intern Med [Internet]. 2000 Sep [cited 2021 Jul 26];15(9):611–9. Available from: http://link.springer.com/10.1046/j.1525-1497.2000.06139.x

50. Pimouguet C, Rizzuto D, Lagergren M, Fratiglioni L, Xu W. Living alone and unplanned hospitalizations among older adults: a population-based longitudinal study. Eur J Public Health [Internet]. 2016 Oct 16 [cited 2021 Jul 26];ckw150. Available from: https://academic.oup.com/eurpub/article-lookup/doi/10.1093/eurpub/ckw150

51. Dreyer K, Steventon A, Fisher R, Deeny SR. The association between living alone and health care utilisation in older adults: a retrospective cohort study of electronic health records from a London general practice. BMC Geriatr [Internet]. 2018 Dec [cited 2021 Jul 26];18(1):269. Available from: https://bmcgeriatr.biomedcentral.com/articles/10.1186/s12877-018-0939-4

52. Bu F, Philip K, Fancourt D. Social isolation and loneliness as risk factors for hospital admissions for respiratory disease among older adults. Thorax [Internet]. 2020 Jul [cited 2021 Jul 26];75(7):597–9. Available from: https://thorax.bmj.com/lookup/doi/10.1136/thoraxjnl-2019-214445

53. Bu F, Abell J, Zaninotto P, Fancourt D. A longitudinal analysis of loneliness, social isolation and falls amongst older people in England. Sci Rep [Internet]. 2020 Dec [cited 2021 Jul 26];10(1):20064. Available from: http://www.nature.com/articles/s41598-020-77104-z

54. Giridhar G, Sathyanarayana KM, Kumar S, James KS, Alam M, editors. Population Ageing in India [Internet]. Cambridge: Cambridge University Press; 2014 [cited 2021 Jul 26]. Available from: http://ebooks.cambridge.org/ref/id/CBO9781139683456

55. United Nations Population Fund. Caring for Our Elders: Early Responses-India Ageing Report 2017 [Internet]. New Delhi, India: UNFPA; 2017 [cited 2021 Jun 30]. Available from: https://india.unfpa.org/sites/default/files/pub-pdf/India%20Ageing%20Report%20-%202017%20%28Final%20Version%29.pdf

56. Banerjee S. Determinants of rural-urban differential in healthcare utilization among the elderly population in India. BMC Public Health [Internet]. 2021 Dec [cited 2021 Jul 26];21(1):939. Available from: https://bmcpublichealth.biomedcentral.com/articles/10.1186/s12889-021-10773-1

57. Sengupta R, Rooj D. The effect of health insurance on hospitalization: Identification of adverse selection, moral hazard and the vulnerable population in the Indian healthcare market. World Development [Internet]. 2019 Oct [cited 2021 Jul 26];122:110–29. Available from: https://linkinghub.elsevier.com/retrieve/pii/S0305750X19301263

58. Sriram S, Khan MM. Effect of health insurance program for the poor on out-of-pocket inpatient care cost in India: evidence from a nationally representative cross-sectional survey. BMC Health Serv Res [Internet]. 2020 Dec [cited 2021 Jul 26];20(1):839. Available from: https://bmchealthservres.biomedcentral.com/articles/10.1186/s12913-020-05692-7

59. Sahoo H, Govil D, James KS, Prasad RD. Health issues, health care utilization and health care expenditure among elderly in India: Thematic review of literature. Aging and Health Research [Internet]. 2021 Jun [cited 2021 Jul 26];1(2):100012. Available from: https://linkinghub.elsevier.com/retrieve/pii/S266703212100010X

60. Bordin D, Cabral LPA, Fadel CB, Santos CB dos, Grden CRB. Factors associated with the hospitalization of the elderly: a national study. Rev bras geriatr gerontol [Internet]. 2018 Aug [cited 2021 Jul 26];21(4):439–46. Available from: http://www.scielo.br/scielo.php?script=sci_arttext&pid=S1809-98232018000400439&lng=en&tlng=en

61. Medeiros SM, Silva LSR, Carneiro JA, Ramos GCF, Barbosa ATF, Caldeira AP. Fatores associados à autopercepção negativa da saúde entre idosos não institucionalizados de Montes Claros, Brasil. Ciênc saúde coletiva [Internet]. 2016 Nov [cited 2021 Jul 26];21(11):3377–86. Available from: http://www.scielo.br/scielo.php?script=sci_arttext&pid=S1413-81232016001103377&lng=pt&tlng=pt

62. Antonelli-Incalzi R, Ancona C, Forastiere F, Belleudi V, Corsonello A, Perucci CA. Socioeconomic status and hospitalization in the very old: a retrospective study. BMC Public Health [Internet]. 2007 Dec [cited 2021 Jul 26];7(1):227. Available from: http://bmcpublichealth.biomedcentral.com/articles/10.1186/1471-2458-7-227

63. Kanitkar SA, Kalyan M, Gaikwad A, Deshmukh S, Saha R. Prevalence of Non Communicable Diseases in Elderly. JIAG [Internet]. 2018 [cited 2021 Jul 27];14(3). Available from: https://www.jiag.org/jiagpdf/2_108_112

64. Figueiredo AEB, Ceccon RF, Figueiredo JHC. Doenças crônicas não transmissíveis e suas implicações na vida de idosos dependentes. Ciênc saúde coletiva [Internet]. 2021 Jan [cited 2021 Jul 27];26(1):77–88. Available from: http://www.scielo.br/scielo.php?script=sci_arttext&pid=S1413-81232021000100077&tlng=pt

65. Yong J, Yang O. Does socioeconomic status affect hospital utilization and health outcomes of chronic disease patients? Eur J Health Econ [Internet]. 2021 Mar [cited 2021 Jul 26];22(2):329–39. Available from: http://link.springer.com/10.1007/s10198-020-01255-z

66. Akinyemi RO, Izzeldin IMH, Dotchin C, Gray WK, Adeniji O, Seidi OA, et al. Contribution of Noncommunicable Diseases to Medical Admissions of Elderly Adults in Africa: A Prospective, Cross-Sectional Study in Nigeria, Sudan, and Tanzania. J Am Geriatr Soc [Internet]. 2014 Aug [cited 2021 Jul 27];62(8):1460–6. Available from: https://onlinelibrary.wiley.com/doi/10.1111/jgs.12940

67. Feng L, Li P, Wang X, Hu Z, Ma Y, Tang W, et al. Distribution and Determinants of Non Communicable Diseases among Elderly Uyghur Ethnic Group in Xinjiang, China. Schnabel RB, editor. PLoS ONE [Internet]. 2014 Aug 20 [cited 2021 Jul 27];9(8):e105536. Available from: https://dx.plos.org/10.1371/journal.pone.0105536

68. Mini GK, Thankappan KR. Pattern, correlates and implications of non-communicable disease multimorbidity among older adults in selected Indian states: a cross-sectional study. BMJ Open [Internet]. 2017 Mar [cited 2021 Jul 27];7(3):e013529. Available from: https://bmjopen.bmj.com/lookup/doi/10.1136/bmjopen-2016-013529

69. Gong JB, Yu XW, Yi XR, Wang CH, Tuo XP. Epidemiology of chronic noncommunicable diseases and evaluation of life quality in elderly. Aging Med [Internet]. 2018 Jun [cited 2021 Jul 27];1(1):64–6. Available from: https://onlinelibrary.wiley.com/doi/10.1002/agm2.12009

70. Banerjee D, Dhar M, Pathania M, Ravikant Rathaur V. Mortality pattern of elderly patients at a tertiary care hospital: A study from Sub-Himalayan region, Uttarakhand, India. J Family Med Prim Care [Internet]. 2019 [cited 2021 Jun 30];8(2):426. Available from: http://www.jfmpc.com/text.asp?2019/8/2/426/253036

71. Swe EE, Htet KKK, Thekkur P, Aung LL, Aye LL, Myint T. Increasing trends in admissions due to non-communicable diseases over 2012 to 2017: findings from three large cities in Myanmar. Trop Med Health [Internet]. 2020 Dec [cited 2021 Jul 27];48(1):24. Available from: https://tropmedhealth.biomedcentral.com/articles/10.1186/s41182-020-00209-8

72. Rahman MM, Kabir MA, Mehjabin M. Pattern of Non-Communicable Diseases among the Admitted Patients in a District Level Hospital of Bangladesh. Bangladesh Heart J [Internet]. 2019 Dec 12 [cited 2021 Jul 27];34(2):118–21. Available from: https://banglajol.info/index.php/BHJ/article/view/44442

73. Kaur G, Bansal R, Anand T, Kumar A, Singh J. Morbidity profile of noncommunicable diseases among elderly in a city in North India. Clinical Epidemiology and Global Health [Internet]. 2019 Mar [cited 2021 Jul 27];7(1):29–34. Available from: https://linkinghub.elsevier.com/retrieve/pii/S2213398417301136

74. Gupta SK, Mahima Mishra BN, Kumar S, Krishnappa K, Shukla SK. Burden of Non-Communicable Diseases at a Tertiary Care Hospital of Central Uttar-Pradesh: A Retrospective Study. OJPM [Internet]. 2018 [cited 2021 Jun 30];08(04):102–8. Available from: http://www.scirp.org/journal/doi.aspx?DOI=10.4236/ojpm.2018.84010

75. Löfqvist T, Burström B, Walander A, Ljung R. Inequalities in avoidable hospitalisation by area income and the role of individual characteristics: a population-based register study in Stockholm County, Sweden. BMJ Qual Saf [Internet]. 2014 Mar [cited 2021 Jul 27];23(3):206–14. Available from: https://qualitysafety.bmj.com/lookup/doi/10.1136/bmjqs-2012-001715

76. Gondek D, Ploubidis GB, Hossin MZ, Gao M, Bann D, Koupil I. Inequality in hospitalization due to non-communicable diseases in Sweden: Age-cohort analysis of the Uppsala Birth Cohort Multigenerational Study. SSM - Population Health [Internet]. 2021 Mar [cited 2021 Jul 27];13:100741. Available from: https://linkinghub.elsevier.com/retrieve/pii/S2352827321000161

77. Taylor CB, Ahn D, Winkleby MA. Neighborhood and Individual Socioeconomic Determinants of Hospitalization. American Journal of Preventive Medicine [Internet]. 2006 Aug [cited 2021 Jul 27];31(2):127–34. Available from: https://linkinghub.elsevier.com/retrieve/pii/S0749379706001632

78. Brameld KJ, Holman CDJ. The use of end-quintile comparisons to identify under-servicing of the poor and over-servicing of the rich: A longitudinal study describing the effect of socioeconomic status on healthcare. BMC Health Serv Res [Internet]. 2005 Dec [cited 2021 Jul 27];5(1):61. Available from: http://bmchealthservres.biomedcentral.com/articles/10.1186/1472-6963-5-61

79. Barnett R, Lauer G. Urban deprivation and public hospital admissions in Christchurch, New Zealand, 1990-1997: Urban deprivation and hospital admissions. Health & Social Care in the Community [Internet]. 2003 Jul [cited 2021 Jul 27];11(4):299–313. Available from: http://doi.wiley.com/10.1046/j.1365-2524.2003.00425.x

80. Joe W, Kumar A, Mishra US. Elderly Inpatient Care Utilization and Financing in India: Is There a Gender Difference? In: Samanta T, editor. Cross-Cultural and Cross-Disciplinary Perspectives in Social Gerontology [Internet]. Singapore: Springer Singapore; 2017 [cited 2021 Jul 1]. p. 245–70. Available from: http://link.springer.com/10.1007/978-981-10-1654-7_13

81. Kastor A, Mohanty SK. Disease-specific out-of-pocket and catastrophic health expenditure on hospitalization in India: Do Indian households face distress health financing? Dwivedi R, editor. PLoS ONE [Internet]. 2018 May 10 [cited 2021 Jun 24];13(5):e0196106. Available from: https://dx.plos.org/10.1371/journal.pone.0196106

82. Shobhana R, Rama Rao P, Lavanya A, Williams R, Vijay V, Ramachandran A. Expenditure on health care incurred by diabetic subjects in a developing country — a study from southern India. Diabetes Research and Clinical Practice [Internet]. 2000 Apr [cited 2021 Jul 1];48(1):37–42. Available from: https://linkinghub.elsevier.com/retrieve/pii/S0168822799001308

83. Tripathy JP, Prasad BM, Shewade HD, Kumar AMV, Zachariah R, Chadha S, et al. Cost of hospitalisation for non-communicable diseases in India: are we pro-poor? Trop Med Int Health [Internet]. 2016 Aug [cited 2021 Jul 1];21(8):1019–28. Available from: https://onlinelibrary.wiley.com/doi/10.1111/tmi.12732

