## Supplementary material for "Differentials and predictors of hospitalization among the elderly people in India: Evidence from 75^th^ round of National Sample Survey (2017-18)": Table S1: Supporting informations.docx

**Table S1** Showing the reported diagnosis and/or main symptoms of the ailments included in the 75^th^ round of NSSO report.

| **Communicable Diseases** | **Non-communicable diseases** | **Injuries & Others ailments** |
| --- | --- | --- |
| Fever with loss of consciousness or altered 01 consciousness, Malaria, Fever due to diphtheria, whooping cough, HIV/AIDS, Other sexually transmitted diseases, Jaundice, Diarrheas/ dysentery/ increased frequency of stools with or without blood and mucus in stools, Worms infestation, Diseases of mouth/teeth/gums, Pain in abdomen: Gastric and peptic, ulcers/ acid reflux/ acute abdomen, Lump or fluid in abdomen or scrotum, Gastrointestinal bleeding, Any difficulty or abnormality in urination, Pain the pelvic region/reproductive tract, infection/ Pain in male genital area. | Cancers (known or suspected by a physician) and occurrence of any growing painless lump in the body, Anaemia (any cause), Bleeding disorders, diabetes, Under-nutrition, Goitre and other diseases of the thyroid, Others (including obesity), Discomfort/pain in the eye with redness or swellings/ boils, Cataract, Glaucoma, Decreased vision (chronic) NOT including where decreased vision is corrected with glasses, Others (including disorders of eye movements – strabismus, nystagmus, ptosis and adnexa), Earache with discharge/bleeding from ear/ infections, Decreased hearing or loss of hearing, Hypertension, Heart disease: Chest pain, breathlessness, Skin infection (boil, abscess, itching) and other skin disease, Joint or bone disease/ pain or swelling in any of the joints, or swelling or pus from the bones, Back or body aches. | Accidental injury, road traffic accidents and falls, Accidental drowning and submersion, Burns and corrosions Poisoning, Intentional self-harm Assault, Contact with venomous/harm-causing animals and plants, Symptom not fitting into any of above categories, Could not even state the main symptom |
